# Craniofacial phenotyping with fetal MRI: A feasibility study of 3D visualisation, segmentation, surface-rendered and physical models

**DOI:** 10.1101/2023.11.30.23299119

**Authors:** Jacqueline Matthew, Alena Uus, Leah De Souza, Robert Wright, Abi Fukami-Gartner, Gema Priego, Carlos Saija, Maria Deprez, Alexia Egloff Collado, Jana Hutter, Lisa Story, Christina Malamateniou, Kawal Rhode, Jo Hajnal, Mary A. Rutherford

## Abstract

This study explores the potential of 3D Slice-to-Volume Registration (SVR) motion-corrected fetal MRI for craniofacial assessment, traditionally used only for fetal brain analysis. In addition, we present the first description of an automated pipeline based on 3D Attention UNet trained for 3D fetal MRI craniofacial segmentation, followed by surface refinement. Results of 3D printing of selected models are also presented.

Qualitative analysis of multiplanar volumes based on the SVR output and surface segmentations outputs, assessed with computer and printed models, were based on standardised protocols that we developed for evaluating image quality and visibility of diagnostic craniofacial features. A test set of 25, postnatally confirmed, Trisomy 21 fetal cases (24-36 weeks gestational age), revealed that 3D reconstructed T2 SVR images provided 66-100% visibility of relevant craniofacial and head structures in the SVR output, and 20-100% and 60-90% anatomical visibility was seen for the baseline and refined 3D computer surface model outputs respectively. Furthermore, 12 of 25 cases, 48%, of refined surface models demonstrated good or excellent overall quality with a further 9 cases, 36% demonstrating moderate quality to include facial, scalp and external ears. Additional 3D printing of 12 physical real-size models (20-36 weeks gestational age) revealed good/excellent overall quality in all cases and distinguishable features between healthy control cases and cases with confirmed anomalies, with only minor manual adjustments required before 3D printing.

Despite varying image quality and data heterogeneity, 3D T2w SVR reconstructions and models provided sufficient resolution for the subjective characterisation of subtle craniofacial features. We also contributed a publicly accessible online 3D T2w MRI atlas of the fetal head, validated for accurate representation of normal fetal anatomy.

Future research will focus on quantitative analysis, optimizing the pipeline, and exploring diagnostic, counselling, and educational applications in fetal craniofacial assessment.

## 1 Introduction

Limitations in 2D and 3D prenatal imaging techniques precludes the reliable assessment of complex cranial and facial structures.[1] Yet, comprehensive prenatal craniofacial assessment is important because, in the setting of an isolated fetal anomaly, an additional craniofacial finding may increase the suspicion of an underlying chromosomal or syndromic condition due to the common association of craniofacial differences [2, 3]. 3D ultrasound (US) is the modality of choice for in-vivo prenatal assessment of superficial facial structures, but its clinical success is limited by fetal position, fetal motion, maternal adiposity, a restricted field of view and overlying tissue or body structures [4]. Magnetic Resonance Imaging (MRI) as a complementary non-ionising fetal imaging technique that is less affected by these limitations, and conventional 2D T2-weighted single-shot turbo spin-echo (SSTSE) sequences are employed for high-risk cases when imaging the fetal brain [5].

### MRI of fetal craniofacial features

Fetal MRI is well established in diagnosis of the fetal brain and body anomalies [6] as well as the characterisation of the normal development patterns. It provides superior tissue contrast and, whilst conventionally 2D MRI methods are involved, 3D fetal MRI methodologies are evolving to complement routine antenatal US. Yet, apart from several narrative and pictorial reviews [7–9], there has been only a limited number of dedicated original studies focusing specifically on fetal MRI of craniofacial features. Zemet (2020), Gai (2022), Arangio (2013) and co-authors, confirmed the added diagnostic value of fetal MRI for evaluation of fetal craniofacial anomalies in retrospective studies comparing MRI and US [10–12]. Other studies have focussed on the MRI imaging of specific features, pathology and measurements within the craniofacial anatomy, for example; the orbits [13–15]; orofacial clefts, including cleft lip and palate [16–21]; inner, middle and external ear structures[22–25]; the upper and lower jaw [26–28]; and skull shape deformities to include craniosynostosis [29–31]. Due to the relative rarity of craniofacial malformations, most MRI studies are retrospective in nature, consist of case series and case studies, and there is a lack of control subjects to assess diagnostic accuracy in a clinical setting.

Notably, there is still a lack of consensus about the methodology for a comprehensive assessment of craniofacial features with fetal MRI, and less regarding methodologies that employ 3D MRI acquisitions, reconstructions, or surface rendering techniques that could complement recommended 3D US imaging protocols [4]. Previously, authors have suggested that the development of three-dimensional surface imaging techniques may broaden the application and effectiveness of fetal MRI in non-central nervous system anatomical areas such as the face [32].

### 3D processing tools for fetal MRI

While T2w MRI provides true 3D spatial information in the acquisition plane and high contrast of the fetal head structures, unpredictable fetal motion remains one of the primary limiting factors since it results in the loss of 3D structural continuity between slices. This is especially critical for biometry measurements that require precise alignment within a 2D image plane, and fetal motion means this cannot be guaranteed during the sequence planning. In the past decade, this challenge has been addressed by retrospective motion correction performed in the image domain [33]. These methods are based on a combination of slice-to-volume registration (SVR) and super-resolution reconstruction that allow 3D reconstruction of high-resolution isotropic images from multiple motion-corrupted stacks of 2D slices. In addition to detailed visualisation, one of the main advantages of SVR reconstructed 3D images is that they can be visualised as multi-planar reformatted (MPR) images in any plane for biometry measurements [34] and segmented to produce 3D volumes of individual structures.

3D SVR reconstructions have been already extensively used in fetal brain MRI research [35] including the development of advanced deep learning methods for multilabel segmentation [36–38]. There have been several works that used deep learning for segmentation of the fetal orbits [39] and preliminary work showing the feasibility of 3D rendering of manual segmentations of fetal craniofacial features from manually segmented 3D images [40, 41]. However, to the best of our knowledge, there have been no large-scale studies that investigate the application of 3D multiplanar SVR images for detailed assessment of the superficial structures of the face and the deep viscerocranium.

Furthermore, automated 3D segmentation of the fetal face has been explored for ultrasound [42], but it has not yet been applied to fetal MRI. This omission is likely due to the detailed and time-consuming manual editing required for high-quality segmentations, the poor differentiation between maternal and fetal tissues, and the limited number of large imaging datasets of homogeneous acquisition types needed to train automated pipelines. In addition, as the manual segmentations are typically performed in 2D planes from the 3D SVR image or raw stacks, with errors between slices and fine surface detail in the region of interest (ROI) not accurately reflecting the tissue interface.

### 3D printing for fetal imaging

In recent years there have been improvements in 3D printing, increased availability, lower costs, and development of biomaterials that have seen the rise in its use in a wide range of healthcare applications [43]. Due to the availability of 3D datasets in radiology, this field has become a primary adopter of the technology, and applications relating to fetal diagnosis and screening have been an emerging area of exploration in recent years [44–47]. The advantages of 3D printing for prenatal craniofacial assessment primarily are for clinician education, parental counselling and education, diagnosis [48] and surgical planning.

Compared to US, MRI data is less sensitive to overlying fetal parts like limbs and cord, and has the advantage of a large ROI covering the whole fetal head and craniofacial region and even body with volume imaging or motion-corrected SVR techniques. However, to date, the literature about high-resolution facial 3D printed models has been based primarily on 3D ultrasound with its limited field of view or true 3D MRI plus 3D US fusion imaging, which may be limited by motion and requires time-consuming manual segmentation techniques [49].

### Contributions

This work provides the first feasibility study for the application of 3D motion-corrected whole-head fetal MRI in the assessment of craniofacial features with respect to both visualisation of diagnostically relevant information in T2w images and the application of automated 3D surface-based analysis. Following the formalisation of protocols for the assessment of general image quality and visibility of diagnostic craniofacial features (deep internal and superficial), we performed a detailed qualitative evaluation on 25 datasets from the Down Syndrome or Trisomy 21 (T21) cohort with different acquisition parameters and gestational age (GA) ranges. Fetuses with T21 are known to have subtle characteristic ‘gestalt’ facial appearances, usually appreciated at birth, but they are rarely qualitatively described prenatally. In addition, the assessment of quality was performed on 12 life-size 3D printed models produced from the 3D reconstructed surfaces of healthy control fetuses and fetuses with confirmed craniofacial anomalies or dysmorphic features related to chromosomal or genetic syndromes (20 - 35 weeks GA range).

As a part of the study, we trained and evaluated the first fully automated pipeline for 3D fetal head segmentation (for T2w MRI) and surface extraction that was then used for the segmentation of the investigated cases. In addition, we created a population-averaged 3D T2w MRI atlas of the fetal head from a set of healthy control subjects for public release and educational purposes.

## 2 Results

### 2.1 Qualitative evaluation of anatomical craniofacial features in 3D MRI SVR

23 anatomical features in the 3D T2w SVR images (n = 25) were assessed for diagnostic visualisation, see fig. 1. 100% visibility was seen for all 11 structures except; the nasal bone (n=16, 66% visibility); the body of the hyoid bone (n = 17, 68% visibility); the body of the mentum (n=19, 76% visibility); the optic nerve (n=23, 92% visibility); and, both external ears (n=19, 76% visibility). The main limitations precluding visualisation were motion-related blurring and poor contrast resolution or ROI adjacent to maternal tissue or extra-cranial fetal structures, see Fig. 3.

**Fig. 1.**
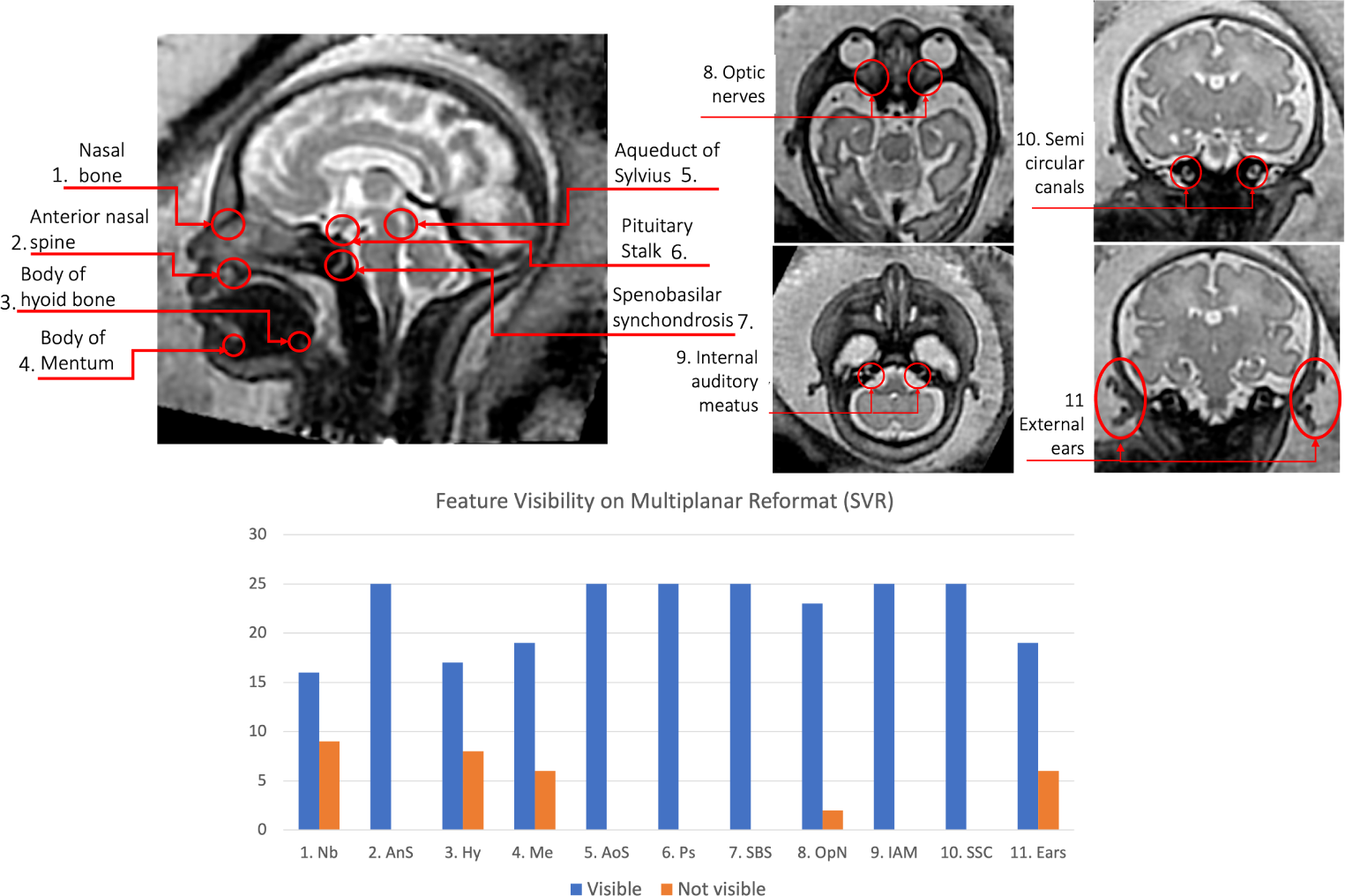
Qualitative features assessed as clear visibility or unclear visibility in SVR images. ‘Nb’ - nasal bone; ‘AnS’ - anterior nasal spine; ‘Hy’ - body of hyoid bone; ‘Me’ - body of mentum; ‘AoS’ - aqueduct of sylvius; ‘Ps’ - pituitary stalk; ‘SBS’ - spenobasilar synchondrosis; ‘OpN’ - optic nerves; ‘IAM’ - internal auditory meatus; ‘SCC’ - semi circular canals; ‘Ears’ - external ears

**Fig. 2.**
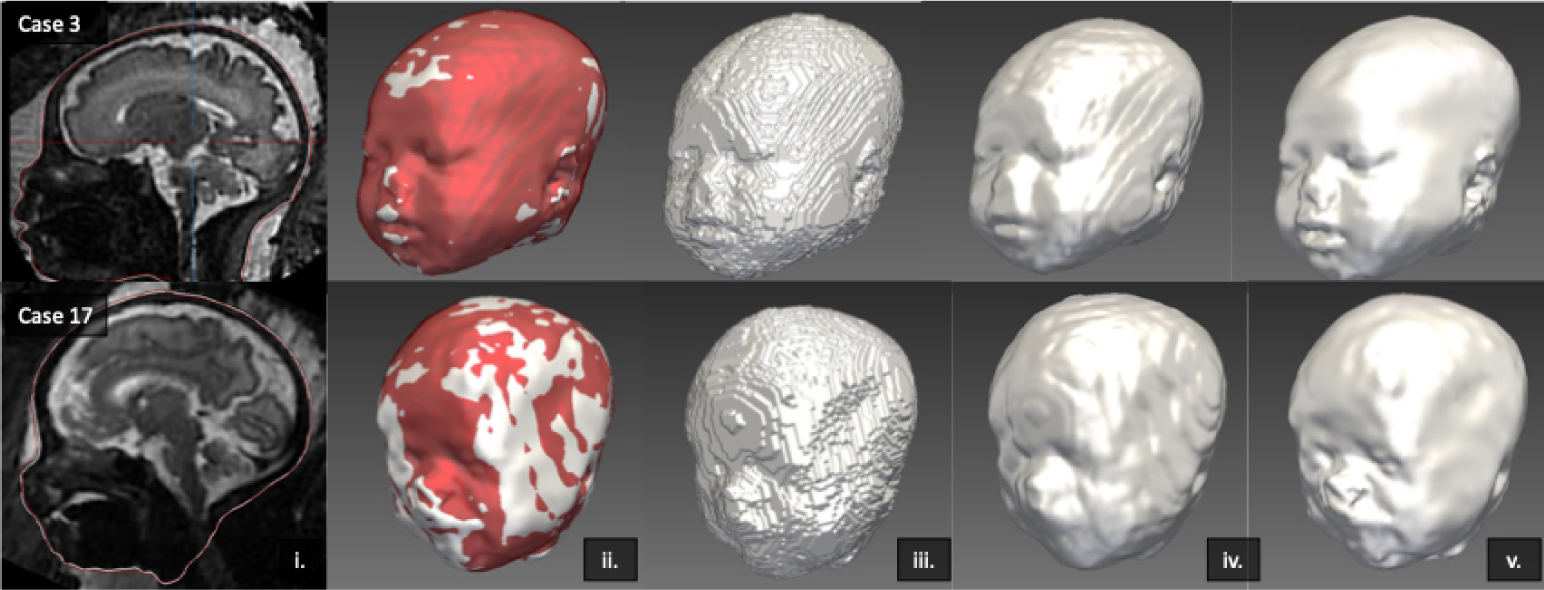
Surface refinement examples of Case 3 and 17 with ‘good’ and ‘moderate’ quality respectively. i) Surface representation over SVR image of baseline output (red) and refined output (white), ii) 3D model overlay of baseline (red) and refined (white) outputs, iii) initial 3D baseline output, iv) smooth polygon baseline model output, v) Surface refinement 3D model output

**Fig. 3.**
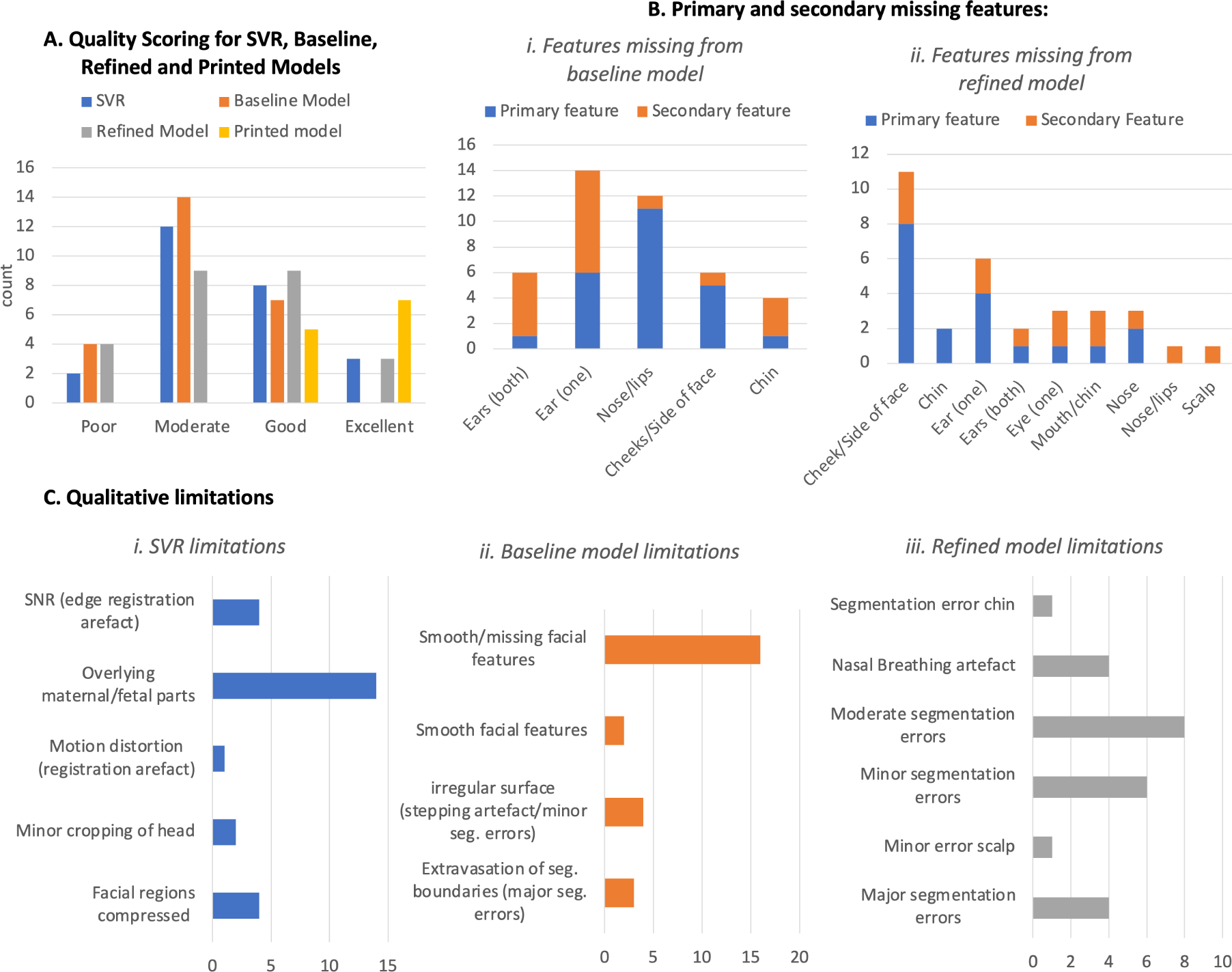
Qualitative evaluation results (SVR images / virtual models n = 25; printed models n = 12): A. Overall quality scoring; B. Primary missing features from, i) baseline and ii) refined models; Q. Qualitative limitations of, i) SVR reconstruction, ii) 3D Attention UNet ‘baseline’ model, and iii) Surface ‘refined’ model

### 2.2 Evaluation of 3D craniofacial surface extraction pipeline

The results of the quantitative evaluation of the network on five previously unseen 3D head images with varying GA, (see table 1), showed good performance when compared to five cases manually segmented in 3D slicer software and resulted in high Dice values (0.970 *±* 0.001), (expected due to the large size of the structure), and were in agreement with recall (0.973 *±* 0.012) and precision (0.967 *±* 0.014). It is also important to note the errors and imperfections in the manual segmentations that were performed in 2D slice-wise.

**Table 1.**
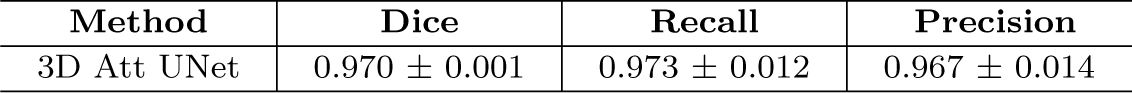
Quantitative evaluation of the 3D face segmentation network on 5 cases with manual segmentation.

| Method | Dice | Recall | Precision |
| --- | --- | --- | --- |
| 3D Att UNet | $0.970 \pm 0.001$ | $0.973 \pm 0.012$ | $0.967 \pm 0.014$ |

While the surface refinement did not change the global segmentation shape, it visibly improved the definition of the finer craniofacial features and smoothed the interpolation errors as shown in Fig. 2.

### 2.3 Qualitative evaluation of fetal craniofacial surface

The results of the global quality and qualitative limitations assessment demonstrated that all 25 SVR images had overall good SNR and included the whole head. However, the SVR demonstrated limitations related to the suitability for surface segmentation. The primary limitation was the proximity of overlying or compressing fetal or maternal tissue to the face/head (e.g. arms, umbilical cord, shoulders, placenta, uterine wall).

Of the 25 cases, the refined model achieved an overall quality score of excellent in three cases (12%) and a good score in nine cases (36%) whereas the baseline model (from the CNN segmentation output) achieved scores of zero excellent and seven (28%) good, with 14 cases being moderate quality (56%).

Errors and omissions in the segmentation were mostly for the fetal ears, nose and lips in the baseline output, whereas when refined, the errors were related to the cheeks, side of the face and one of the ears. The main limitation of the baseline segmentation was ill-defined and smoothed or missing facial features, whereas, for the refined model, segmentation errors were the key issue i.e. moderate extravasation at boundaries or minor irregularities.

When comparing the detailed feature visualisation in a subset of the baseline models and refined models (n = 10), there were finer facial details seen in the refined model for 17 of the 23 structures assessed i.e. the nose, right ear, eyes and lip detail being visualised more consistently (see Fig. 4).

**Fig. 4.**
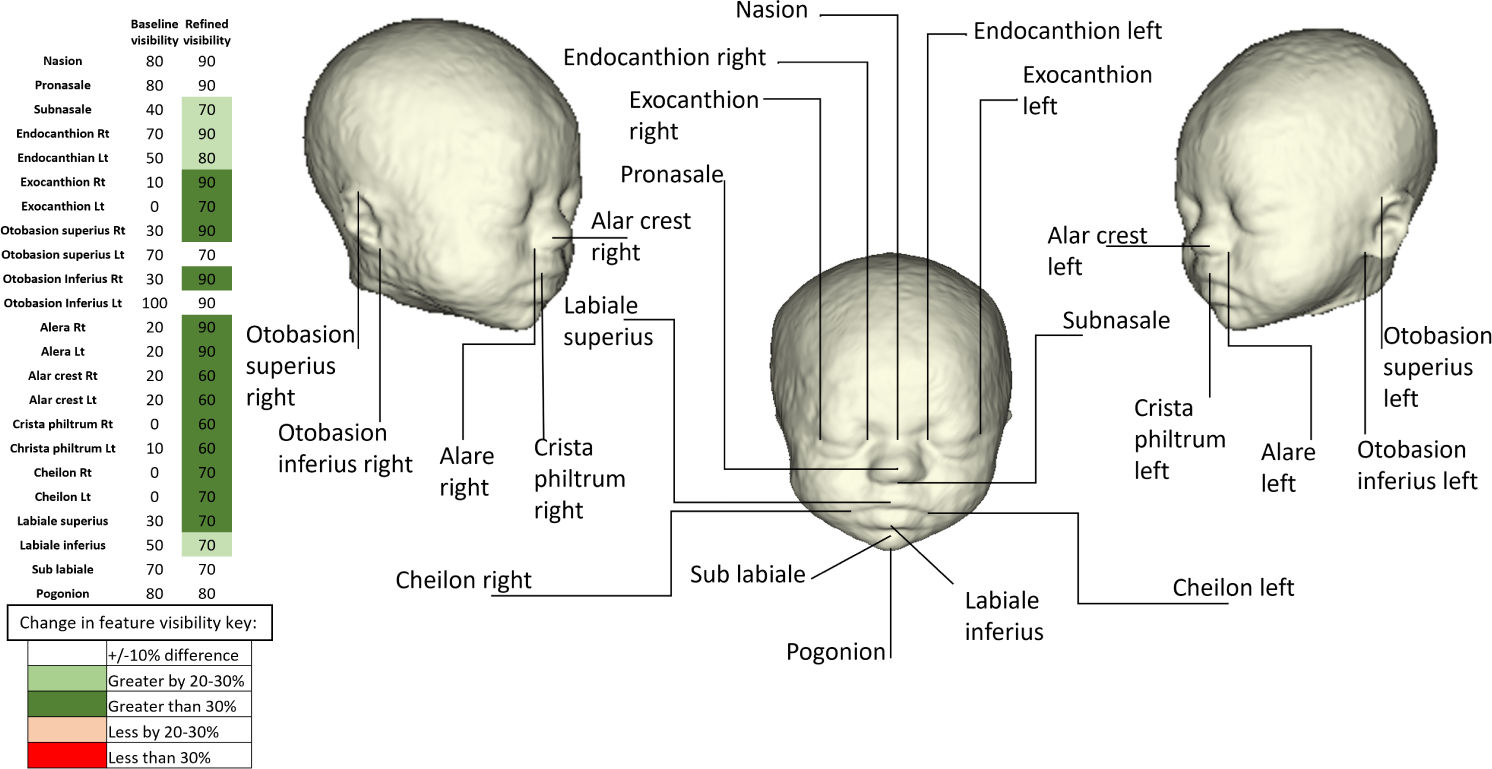
Labelled diagram of anatomical craniofacial landmarks and table of proportion (%) of visualised landmarks across 10 random test cases for the baseline and refined 3D model outputs

### 2.4 3D printing of fetal craniofacial features

All 12 models were successfully printed with a resultant quality score of good (five cases, 42%) or excellent (seven cases, 58%), see Fig. 5-6. A video of all printed models is available in the supplementary files and ‘here’.

**Fig. 5.**
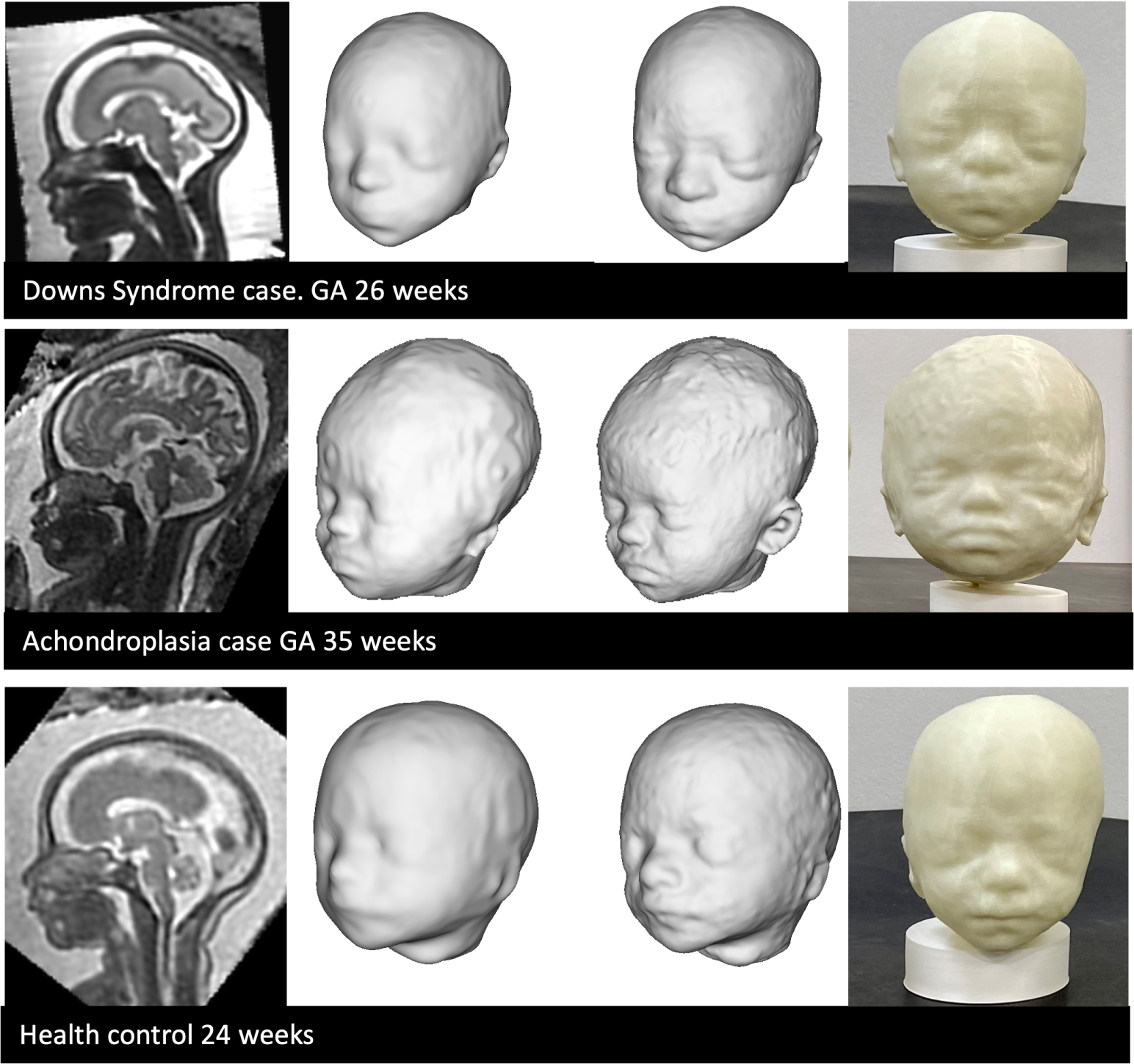
3D printed example results, including (from left to right): SVR image, baseline 3D surface model, refined surface 3D model, printed 3D model

**Fig. 6.**
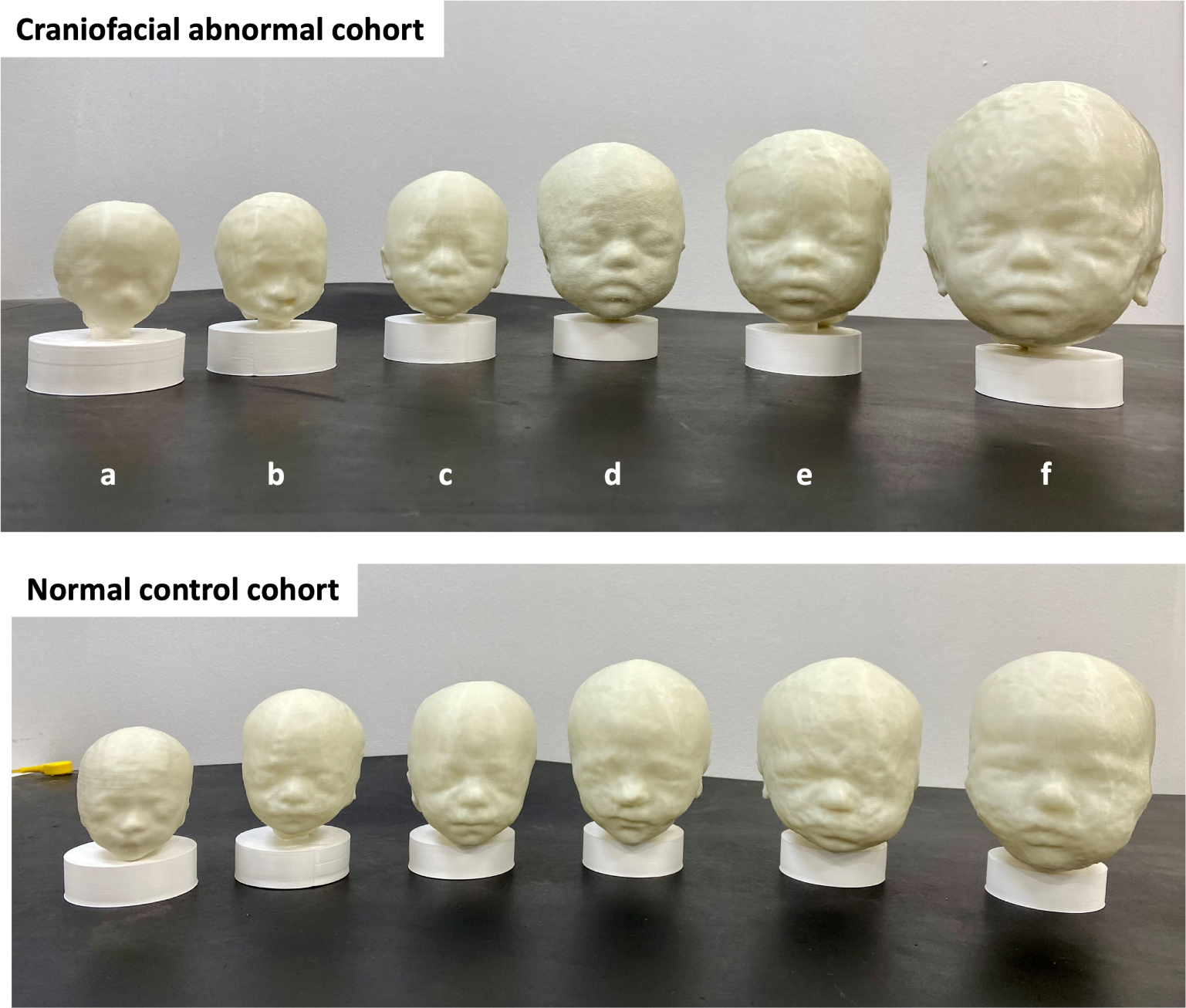
All 3D printed cases from the cohorts with craniofacial anomalies (abnormal) and normal healthy subjects from early to late GA (20 - 35 weeks) in real-life size. Abnormal conditions: a.Trisomy 18; b. Cleft lip/palate; c-e. Trisomy21; and, f. Achondroplasia

The physical models were scored as ‘good’ rather than ‘excellent’ if there were minor irregularities in the surface, and it was noted that this was most commonly seen in the scalp, ear and jaw area, likely due to proximity to the uterine wall and increased noise, due to jaw motion or other overlying structures. In six cases, one ear required manual editing, in three cases both ears, and in three neither ear required edits.

The visual assessment of the physical models in Fig. 6, provided a means of subjectively assessing the gestalt appearance and dysmorphic features or structural facial anomalies. In the high-risk for craniofacial anomaly cohort, features such as upward slanting eyes, down-turned lips, flattened nasal bridge and flattened occiput were identified in cases c, d and e, consistent with the T21 gestalt, however, this was more obvious at the later GAs (28 and 32weeks). Model-a was diagnosed with confirmed Trisomy 18 at 20 weeks GA and appeared to have a small chin but it was difficult to subjectively define distinct craniofacial dysmorphic features from the healthy control early GA model and corresponding facial appearances. Model-b had a left-sided cleft lip which was clearly visible, and Model-f had a diagnosis of achondroplasia, with visual features including frontal bossing, round/large head shape, small midface with flattened nasal bridge.

### 2.5 3D population-averaged fetal head MRI atlas

The population-averaged atlas (Fig. 9.A) was inspected by clinicians, JM and MR, who confirmed that all craniofacial features had clear visibility, were well-defined and physiologically correct, subjectively corresponding to the normal fetal development. The head surface also corresponded to the expected normal appearance of craniofacial features. The 3D printed atlas is shown in Fig. 9.B also includes the separate brain surface model subtracted in order to reveal the base of skull.

## 3 Methods

### 3.1 Cohort, acquisition and pre-processing

Participants were scanned between 2014 and 2023 at a single site (St.Thomas’ Hospital, London, UK) and all maternal participants gave written informed consent for the use of data acquired under one of five MRI research studies: The Placental Imaging Project (PIP, REC 14/LO/1169)^1^; the Intelligent Fetal Imaging and Diagnosis (iFIND, REC 14/LO/1806); the quantification of fetal growth and development with MRI study (fetal MRI, REC 07/H0707/105)^2^; the fetal CMR service at Evelina London Children’s Hospital (REC 07/H0707/105); the Individualised risk prediction of adverse neonatal outcome in pregnancies that deliver preterm using advanced MRI techniques and machine learning study (PRESTO: REC 21/SS/0082); early brain imaging in Down syndrome study (eBiDS, REC 19/LO/0667); and, the fetal imaging with maternal oxygen study (FIMOx, REC 17/LO/0282).

#### 3.1.1 MRI acquisition parameters

The MRI acquisitions across the cohorts were performed at St.Thomas’s Hospital, London on one of 3 MRI machines (Philips Ingenia 1.5T, Philips Achieva 3T, Siemens Sola 1.5T) with 4 different T2-weighted acquisition protocols covering the brain ROI.

#### 3.1.2 Datasets

The automated segmentation pipeline was trained on data from 76 subjects without structural anomalies (with an additional 5 reserved for validation), with two different acquisition protocols (either 1.5T, TE=80ms or 3T, TE=180 ms), at a GA range of 24-38 weeks. The MRI datasets and parameters included:

- 48 datasets acquired on 1.5T Philips Ingenia MRI system using 28-channel torso coil with TE=80ms and TE=180ms, acquisition resolution 1.25x1.25mm, slice thickness 2.5mm, -1.25mm gap and 9-11 stacks;
- 50 datasets acquired on 3T Philips Achieva MRI system using a 32-channel cardiac coil with TE=180ms, acquisition resolution 1.25 x 1.25mm, slice thickness 2.5, - 1.5mm gap and 5-6 stacks;
- 5 datasets acquired on 3T Philips Achieva MRI system with a 32-channel cardiac coil using a dedicated dHCP fetal acquisition protocol with TE=250ms, acquisition resolution 1.1 x 1.1mm, slice thickness 2.2mm, -1.1mm gap and 6 stacks;
- 3 datasets acquired on 1.5T Siemens Sola MRI system using 28-channel torso coil with TE=180ms, acquisition resolution 1.25 x 1.25mm, slice thickness 3mm and 9-11 stacks.

The main test dataset for qualitative assessment was a cohort with confirmed T21 and included 25 subjects scanned at different MRI field strengths and a GA ranging from 24 to 36 weeks. Lastly, a set of 12 fetuses including healthy control subjects and subjects with confirmed craniofacial anomalies were selected between 20 and 36 weeks GA at regular intervals to assess the feasibility of manufacturing physical printed models from the automated pipeline outputs. The 6 cases with craniofacial anomalies included; three cases of T21; one case of achondroplasia, AC; one case of Trisomy 18 (Edwards Syndrome), T18; and, one case of cleft lip and palate. These 3D print cases and the qualitative test datasets were selected from either the iFIND, fetal MRI, PiP, PRESTO, eBiDS or FIMOx studies.

#### 3.1.3 3D SVR head reconstruction

All datasets were reconstructed for the whole head using the optimised version of the classical 3D SVR reconstruction method [50] in SVRTK package^3^ [51] to 0.8mm isotropic resolution and semi-manually reoriented to the standard radiological space (see Fig. 7A). Successful 3D reconstructions were included and defined as containing the full cranial and facial ROI, i.e., the exclusion criteria was insufficient coverage of the face ROI in original stacks. Cases were not excluded where the fetal head was directly adjacent to maternal or extra-cranial fetal parts or for regional suboptimal image quality (e.g., blurring of craniofacial features due to motion).

**Fig. 7.**
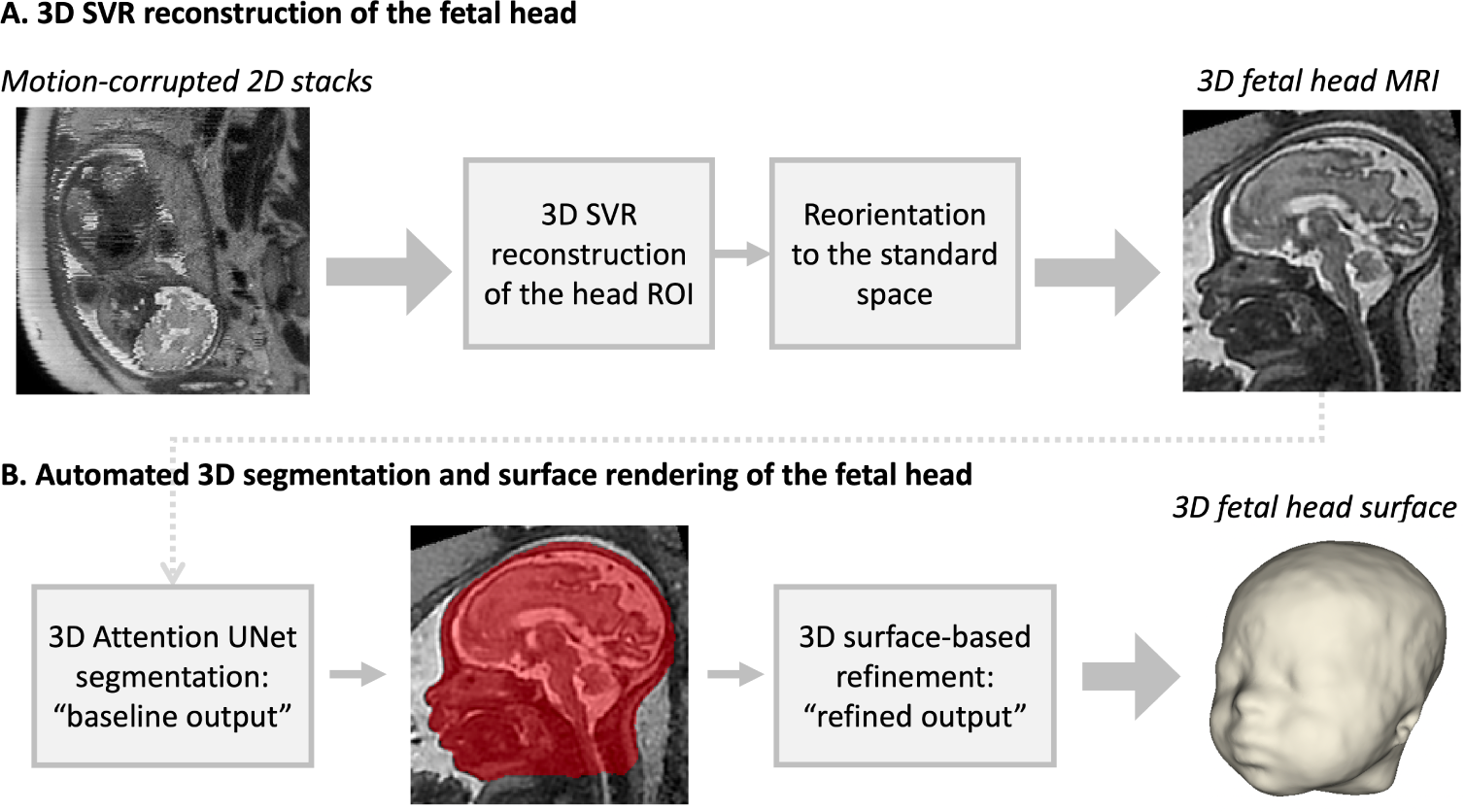
Proposed pipeline for 3D SVR reconstruction (A) and automated face surface extraction (B) for 3D fetal head MRI.)

### 3.2 Qualitative evaluation of detailed craniofacial features in 3D MRI SVR

To assess detailed craniofacial structures within the reconstructed fetal MRI head volume, the multiplanar reformatted images were adjusted to obtain precisely aligned orthogonal planes and 11 subtle structures were reviewed using the 3DSlicer platform^4^ (Massachusetts, USA) [52]. The structures are outlined in figure 1 and included the; nasal bone; anterior nasal spine; body of hyoid bone; body of mentum; aqueduct of sylvius; pituitary stalk; spenobasilar synchondrosis; and bilateral structures of the optic nerves, internal auditory meatus, semicircular canals and the external ears, which were all scored as one item. The features to be assessed were agreed by consensus between two clinicians (fetal neuroradiologist (GP) and obstetric reporting radiographer (JM), both with more than 10 years of experience). The scoring criteria were agreed, based on binary outcomes being: 1. visible (high image quality at a diagnostic level) or 2. not visible (suboptimal visualization for diagnostic interpretation). A reviewer training set of three cases were assessed independently by the clinicians, who then met to discuss any discrepancies. All 25 cases in the T21 test cohort were then scored by a single operator (JM). The global quality assessment of the SVR output is described in section 3.4.

### 3.3 Automated 3D craniofacial surface extraction pipeline

The proposed pipeline for automated 3D surface extraction of the fetal head from 3D motion-corrected fetal head images summarised in Fig. 7B includes deep learning segmentation followed by automated surface-based refinement.

#### 3.3.1 3D segmentation network

For the face segmentation network we used the standard MONAI [53] 3D Attention-UNet [54] implementation with five and four encoder-decoder blocks (output channels 32, 64, 128, 256 and 512), correspondingly, convolution and upsampling kernel size of 3, ReLU activation, dropout ratio of 0.5. We employed AdamW optimiser with a linearly decaying learning rate, initialised at 1*×*10*^−^*^3^, default *β* parameters and weight decay=1 *×* 10*^−^*^5^. The input image dimensions are 128x128x128 and the outputs have 2 channels (head label and background).

Taking into account the varying size, resolution and intensity ranges of input SVR reconstructions, the general preprocessing steps included: transformation to the standard radiological space coordinate system, cropping of the background, resampling with padding to the required input grid size and histogram matching to TE=80ms sample image (to increase the image contrast) followed by rescaling to 0-1. All preprocessing steps were implemented based on MIRTK toolbox^5^.

We used the semi-supervised training strategy in 3 iterations where the training dataset was expanded by manual refinement of the network output from the previous iteration and the final testing was performed in the 5 cases using manual ground truth labels. The final training dataset includes 76 segmented head images.

#### 3.3.2 3D surface based refinement

Segmentation masks were refined using previous approaches related to brain cortical surface refinement, modelling the head as a closed genus-0 surface (spherical topology), which is deformed by “internal” and “external” forces [55–57]. Firstly, a bounding sphere was deformed inward towards the zero level set of the signed Euclidean distance transform of the segmentation. Remeshing at each iterations allows the surface to locally expand or contract and adapt to the head geometry.

The relatively low resolution of the original 3D Attention UNet segmentation fails to accurately capture more detailed features of the face, such as the ears, lips and eyelids. For this reason, a second deformation procedure was used to adapt the surface towards the skin/amniotic fluid contour. A combination of four forces were used: 1) a distance force that ensures the mesh remains close to the original segmentation; 2) a balloon inflation force to expand the model into under-segmented areas (e.g., the ears)[58]; 3) an edge-distance force to snap the mesh to the skin/amniotic fluid contour [56]; and 4) a smoothing force which reduces curvature, avoiding voxelisation [58].

### 3.4 Qualitative evaluation of fetal craniofacial surface extraction pipeline

A qualitative evaluation was performed for the 3 steps of the pipeline in the test dataset: 1. The SVR image reconstruction: 2. The 3D UNet output or ‘baseline’ 3D model: 3. The surface refinement output or ‘refined’ 3D model, using 3DSlicer software.

#### Global quality assessment

For the 25 test subjects and three steps in the pipeline (SVR output, baseline model, refined model), the overall quality was recorded on a scale of 1-4 by a single operator and trained clinician, JM (1=poor, 2=moderate, 3=good, 4=excellent, see Fig. 8 for example scoring).

**Fig. 8.**
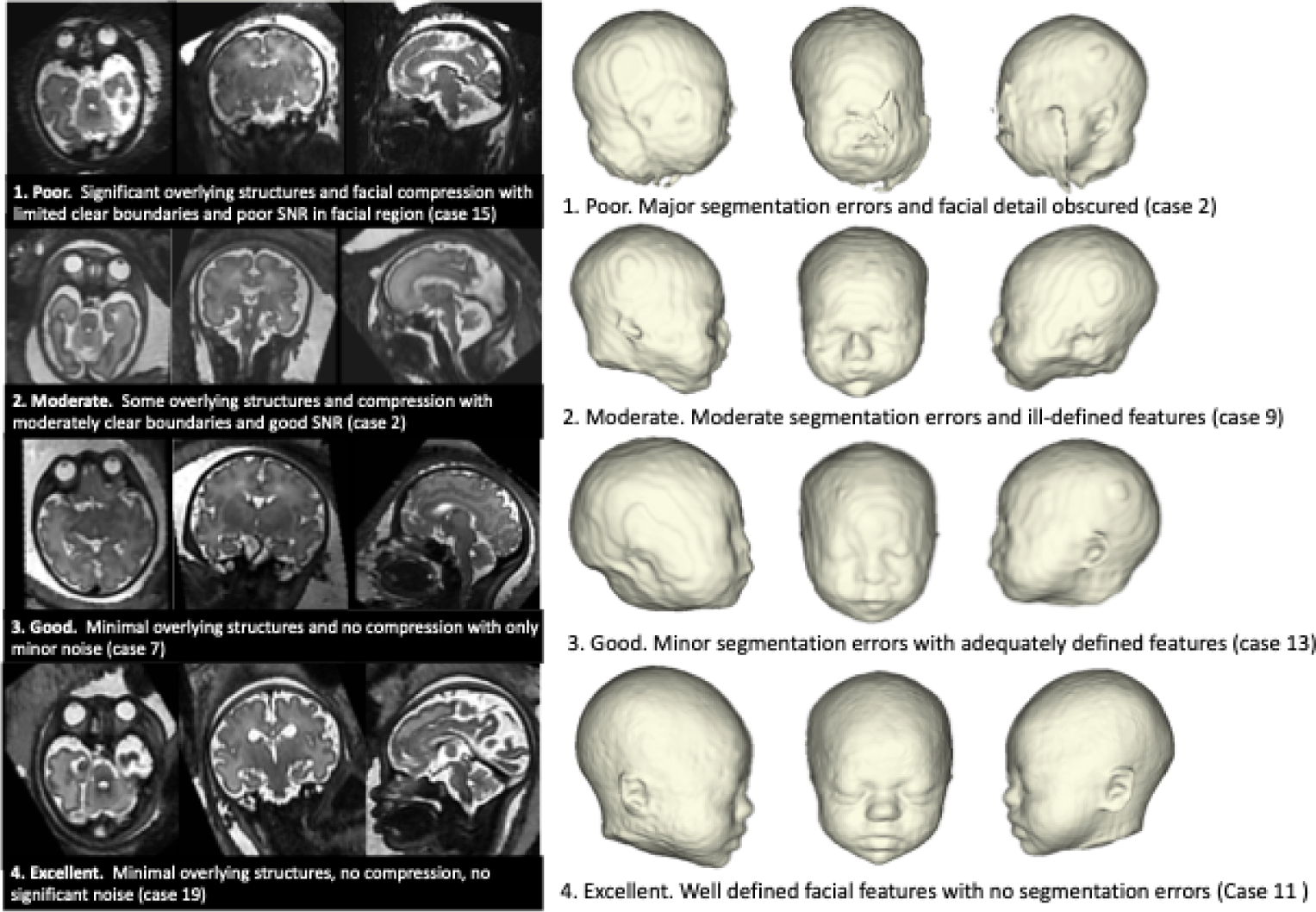
3D SVR image and virtual model quality scoring guide with example images

**Fig. 9.**
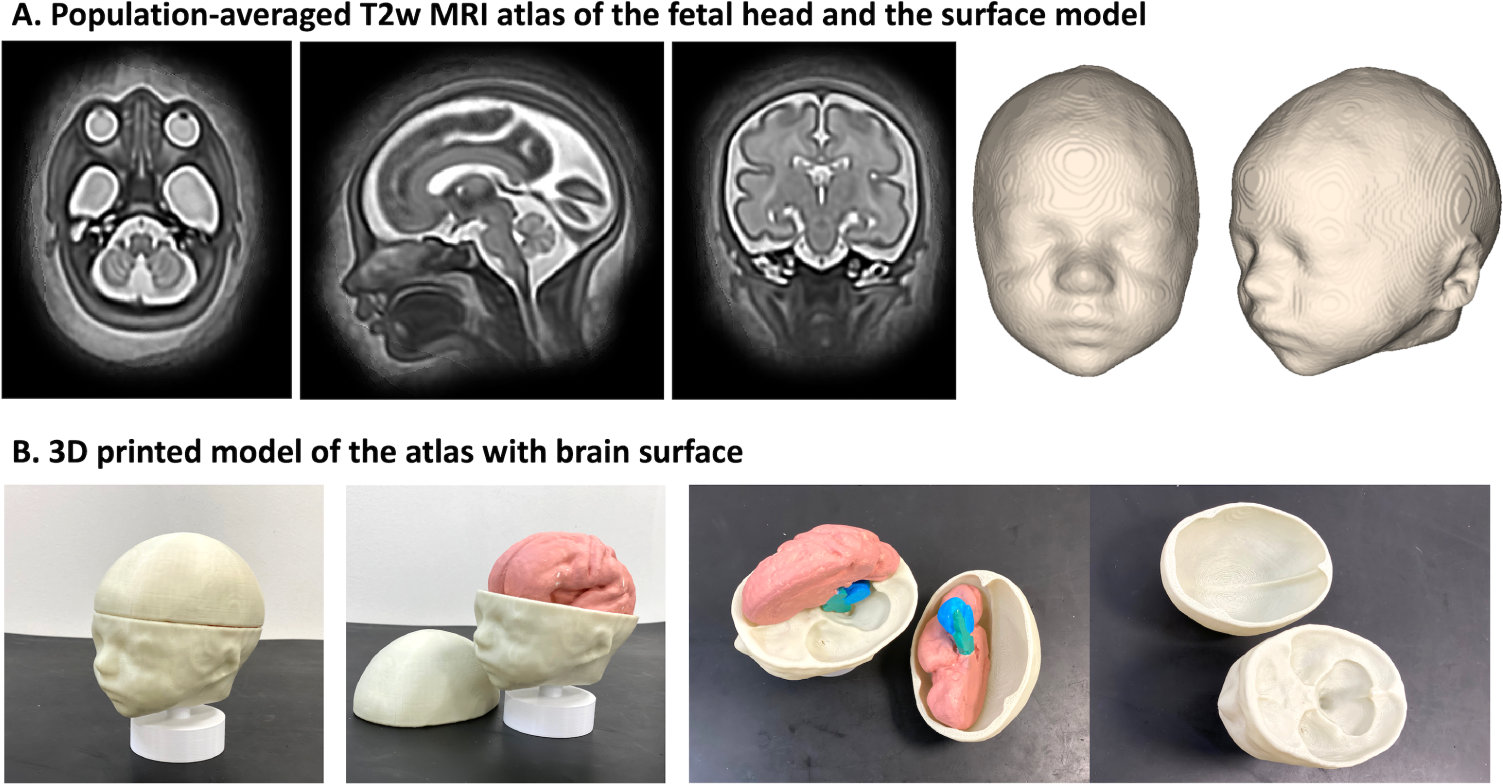
3D population-averaged fetal head MRI atlas and the corresponding surface segmentation (A) and 3D printed physical model (B) with additional brain surface

#### Qualitative limitations assessment

Limitations in the SVR reconstruction that may impact a successful segmentation were recorded, as were the limitations of the baseline 3D UNet output and the refined model output. These limitations were categorised into primary and secondary observations (in order of significance), with no apriori categories provided. Any similar categories were subsequently grouped into single themes once all the data had been collected.

#### Feature visualisation comparison

In a random sample of 10 test cases across the gestational age range, a detailed qualitative evaluation was performed. 23 superficial facial anatomical landmarks, as proposed by Alomar et al. in 2022[59], were reviewed for the baseline and refined 3D virtual surface model, see Fig. 4. Each landmark was scored as either visible or not visible based on whether the observer could accurately and confidently identify the landmark.

### 3.5 3D printing of fetal craniofacial features

For 3D printing, we also used additional minor manual editing of the proposed automated segmentation outputs, Fig. 7, before the surface refinement in order to achieve higher anatomical accuracy and correct minor errors.

A Flsun3D QQ-s (Zhengzhou, Henan, China) 3D printer was used to test the 31-week template atlas, see section 3.6, and included a calvarial cut to aid visualisation of a 3D-printed brain and the inner table of the cranial vault and base. The printing parameters are given in table 2.

**Table 2.** 3D printing parameters used in this study.

| Parameter | Value |
| --- | --- |
| Printer model/manufacturer | Flsun3D QQ-s, (Zhengzhou, Henan, China) |
| Printing process | Fused Deposition Modelling, FDM |
| Filament type | Polylactic Acid Plus, PLA+ |
| Nozzle size | 0.4mm |
| Printer resolution | 0.2mm |
| Layer height | 0.2mm |
| Wall thickness | 0.8mm |
| Infill density | 5% |
| Print speed | 60 mm/s |
| Printing temperature | 215 degrees |
| Built plate temperature | 60 degrees |
| Price range (materials only per case) | 0.3 - 2.1 GBP |
| Time to print (per case) | 1-6 hours |

In the remaining print cases, the manual refinement was performed using 3DSlicer software^6^ and the refinement corrections were focused on the detailed segmentation of the fetal external ears, taking less than 5 minutes per case. On completion of the surface refinement, the models were saved in stereolithography (.stl) format and imported into Cura Slicer software (Ultimaker Ltd., The Netherlands) where the printer parameters were specified. The models were resampled to 0.1mm resolution. As part of the design and manufacturing process, stands and base-plates were developed using Fusion 360 CAD software (Autodesk, California, USA) for accurate model alignment and placement.

Once the prints were completed, the printing supports were removed and the models were smoothed at the locations where supports were attached. At the final step, they were fitted onto the printed cylindrical 2cm tall stands with a 2cm thick cylindrical base 6cm in diameter. The stands included a rectangular cut-out (0.3cm by 1cm, offset of 0.1cm) to allow attachment to the laser-cut base plate.

The quality of the final 12 printed models was assessed based on the quality scoring as described for the virtual model QC protocol in Fig. 8.

### 3.6 3D population-averaged fetal head MRI atlas

The population-averaged atlas of the fetal head was created using MIRTK^7^ *construct-atlas* tool from 12 normal subjects in the iFIND cohort (1.5T, TE=80ms). The inclusion criteria were 29-31 weeks GA, good reconstruction quality, and clear visibility of all craniofacial structures. We used standard settings with local crosscorrelation similarity metric, 3 iterations and 0.8mm resolution. The final atlas was resampled to a 0.5mm grid. We used the trained network to segment the atlas followed by smoothing. The atlas is publicly available online at the KCL CDB data repository^8^.

## 4 Discussion

### Feasibility of 3D MRI fetal models

This work presents the first study of the application of a large ROI 3D SVR motion-corrected fetal MRI. While 3D SVR reconstruction to correct for fetal motion has been widely employed for 3D multiplanar analysis of the fetal brain development and anomalies, the reconstructed images normally omit lower and superficial facial regions. Utilising the presented imaging methodology, assessing craniofacial features with respect to both visualisation of diagnostically relevant information in T2w images and automated 3D surface-based models is feasible.

Firstly, we formalised a set of protocols for qualitative scoring of 3D T2w SVR images and surface models in terms of general image quality and visibility of diagnostic craniofacial features (deep internal and superficial structures). This included assessments of 11 T2w SVR grayscale structures and 23 surface-based craniofacial landmarks and features relevant to diagnostic evaluation.

On implementing an automated deep learning pipeline for the 3D whole fetal head and face segmentation with an additional refinement step for generation of virtual surface models, we demonstrate the feasibility of obtaining a high-quality yield with this pipeline. Our results demonstrate that the classical implementation of 3D Attention UNet provides a sufficient quality baseline for surface-based refinement of finer features and has the potential for practical clinical or research-based assessments that would otherwise require extensive resources related to manual segmentations and time.

We performed a detailed qualitative evaluation on the extracted head surfaces in the 25 datasets from the cohort with Down Syndrome from 24 - 34 weeks gestational (GA) range. The MRI protocols were heterogeneous representing the expected variability of the real-world clinical data. There was only a small proportion of subjects with major segmentation errors and very detailed features could be visualised with MRI 3D surface-rendered models. The superficial features of the ears, eyes, head shape and mouth shape could be easily discerned by a trained clinician, and in this cohort the superficial features were consistent with the T21 gestalt (i.e. subjective facial appearance associated with T21) consistent with subjective superficial anatomic assessment currently possible with ultrasound [4]. Using the same pipeline, we printed 12 3D reconstructed cases in life-size from normal control and abnormal cohorts (20 - 35 weeks GA range). The models demonstrate visible differences between the normal and abnormal cases as well as the expected changes with gestation. Lastly, we created a population-averaged 3D T2w MRI atlas of the fetal head from a set of healthy control subjects. Clinicians experienced in fetal MRI confirmed that the model provides accurate representation of normal fetal anatomy. The atlas is publicly available for both research and educational purposes.

### Applications of 3D MRI craniofacial models

Understanding which features are achievable in this modality is the first step in characterising anatomical landmarks that can be used in advanced facial analysis. 3D morphometric and statistical shape models have been used in the paediatric and adult populations and have an emerging application in studying fetal facial anatomy with 3D ultrasound to study models of variations in anatomical structures [59–61]. This study indicates the feasibility of applying similar methodologies with fetal MRI with the added advantages of comprehensive coverage of the whole face and scalp regions.

With the use of 3D T2w SVR reconstructions and surface-renders, appreciation of individual and subtle changes in facial morphology were possible antenatally although currently they are often detected after birth during a newborn assessment[62]. Even with the given limitations in terms of varying image quality, visibility of superficial features and the heterogeneity of the MRI protocols in the test cases, the images had sufficient resolution and quality to subjectively characterise subtle dysmorphic craniofacial features. Applications include deeper radiological phenotyping of craniofacial features related to genetic or syndromic conditions, parental education after a diagnosis e.g. cleft lip, training of imaging professionals, and there may be a role in parental bonding or for visually impaired parents [63, 64].

### Limitations and future work

In terms of limitations, integration of 3D fetal craniofacial T2w MRI into clinical practice would require further assessment of image quality and visibility of various structures with a wider range of acquisition protocols and types of craniofacial anomalies compared to clinical ground truth. For example, the lower yield of nasal bone structures visualised in the SVR images for T21 cohort is a feature typically noted physiologically and on 2D ultrasound.

Automation of 3D SVR reconstruction and reorientation for the whole head ROI would also be a useful addition to the surface extraction in order to fully automate the proposed analysis pipeline. Although, previous work has focused on the fetal orbits[39], only one binary label map was included and further subdivisions into deep anatomical regions e.g. mandible, and the feasibility of deeper anatomical characterisation with segmented structures may widen clinical applications in the future.

Furthermore, the subjective nature of the quality scoring of the 3D T2w reconstructed images and models will benefit from an inter-observer assessment in future studies to validate findings. In addition, no criteria were set to comment on the limitations within the SVR images or the 3D models, and further assessments with well-defined categories will aid in understanding what characteristics within the SVR volume predict a high-quality 3D model. Likewise, defined categories will help to provide a deeper understanding of the reliability of model features related to the grey-scale image, which is important to understand any artefacts generated by this method.

Whilst this study has focused on the qualitative evaluation of the output similar to current radiological practice, there are opportunities to quantitatively assess fetal craniofacial development [65]. Craniofacial research is an active area of exploration in the neonatal and paediatric patient groups with 3D morphometric and shape analysis performed with 3D imaging, 2D and 3D photogrammetry [66–68] and emerging prenatal methods have been proposed [60].

Future work should focus on development of fetal MRI sequences to image craniofacial bones directly, for example, black bone imaging or zero TE imaging [69, 70]. Further optimisation of the segmentation pipeline to include deeper investigation of possible application areas for intensity- and surface-based analysis and 3D printing including quantification of improvement in diagnostic confidence, parental counselling, as well educational materials.

## 5 Conclusions

In conclusion, this work confirmed the general feasibility of using 3D T2w MRI for detailed assessment of fetal craniofacial anatomy. Furthermore, the production of individualised virtual and physical fetal models in-vivo (from automated fetal MRI segmentations) is realistic and has potential applications for characterising the craniofacial phenotype in screening, diagnostic applications, education and parental counselling in the setting of rare conditions.

## Author Contributions

J.M. and A.U. contributed equally to this work and prepared the manuscript. J.M. analysed the datasets, formalised qualitative evaluation protocols, performed all qualitative evaluation experiments, refined models for 3D printing and prepared all illustrations. A.U. reconstructed 3D SVR images, implemented automated learning component of the segmentation pipeline, processed the datasets and created the 3D atlas. L.D.S. and C.S. optimised the 3D printing protocol and printed the 3D head models. R.W. implemented the automated refinement step for face surface extraction pipeline. A.F.G. participated in processing and analysis of T21 cohort cases. M.D. supervised various parts of the project. G.P. contributed to formalisation of visualisation assessment protocols. A.E.C. contributed to diagnosis of the datasets. J.H. and L.S. provided fetal MRI datasets. C.M. supervised various parts of the project. K.R. supervised 3D printing and provided all materials. J.V.H. provided fetal MRI datasets and supervised various parts of the project. M.A.R. provided fetal MRI dataset, contributed to analysis of the datasets and supervised the project. All authors reviewed the final version of the manuscript.

## Data Availability

The individual fetal MRI datasets used for this study are not publicly available due to ethics regulations. The created average normal 3D T2w atlas is publicly available online at KCL CDB fetal body MRI atlas repository.

https://gin.g-node.org/kcl_cdb/craniofacial_fetal_mri_atlas

## Acknowledgments

We thank everyone who was involved in the acquisition and analysis of the datasets at the Department of Perinatal Imaging and Health at King’s College London and St Thomas’ Hospital. We thank all participants and their families.

The views expressed are those of the authors and not necessarily those of the NHS, the NIHR or the Department of Health.

## Footnotes

1 PiP project: https://placentaimagingproject.org/project/

2 iFIND project: https://www.ifindproject.com/

3 SVRTK toolbox: https://github.com/SVRTK/SVRTK

4 3D Slicer software: www.slicer.org

5 MIRTK toolbox: https://github.com/BioMedIA/MIRTK

6 3D Slicer software: https://www.slicer.org/

7 MIRTK toolbox: https://github.com/BioMedIA/MIRTK

8 KCL CDB atlas repository: https://gin.g-node.org/kcl_cdb/craniofacial_fetal_mri_atlas

